# Genomic epidemiology of *Campylobacter jejuni* associated with asymptomatic pediatric infection in the Peruvian Amazon

**DOI:** 10.1101/2020.04.26.20075689

**Authors:** Ben Pascoe, Francesca Schiaffino, Susan Murray, Guillaume Méric, Sion C. Bayliss, Matthew D. Hitchings, Evangelos Mourkas, Jessica K. Calland, Rosa Burga, Pablo Peñataro Yori, Keith A. Jolley, Kerry K. Cooper, Craig T. Parker, Maribel Paredes Olortegui, Margaret N. Kosek, Samuel K. Sheppard

## Abstract

*Campylobacter* is the leading bacterial cause of gastroenteritis worldwide and its incidence is especially high in low- and middle-income countries (LMIC). Disease epidemiology in LMICs is different compared to high income countries like the USA or in Europe. Children in LMICs commonly have repeated and chronic infections even in the absence of symptoms, which can lead to deficits in early childhood development. In this study, we sequenced and characterized *C. jejuni* (n=62) from a longitudinal cohort study of children under the age of 5 with and without diarrheal symptoms, and contextualized them within a global *C. jejuni* genome collection. Epidemiological differences in disease presentation were reflected in the genomes, specifically by the absence of some of the most common global disease-causing lineages. As in many other countries, poultry-associated strains were a major source of human infection but almost half of local disease cases (15 of 31) were attributable to genotypes that are rare outside of Peru. Asymptomatic infection was not limited to a single (or few) human adapted lineages but resulted from phylogenetically divergent strains suggesting an important role for host factors in the cryptic epidemiology of campylobacteriosis in LMICs.

**Author summary:** *Campylobacter* is the leading bacterial cause of gastroenteritis worldwide and despite high incidence in low- and middle-income countries (LMICs), where infection can be fatal, culture based isolation is rare and the genotypes responsible for disease have not broadly been identified. The epidemiology of disease is different to that in high income countries, where sporadic infection associated with contaminated food consumption typically leads to acute gastroenteritis. In some LMICs infection is endemic among children and common asymptomatic carriage is associated with malnutrition, attenuated growth in early childhood, and poor cognitive and physical development. Here, we sequenced the genomes of isolates sampled from children in the Peruvian Amazon to investigate genotypes associated with varying disease severity and the source of infection. Among the common globally circulating genotypes and local genotypes rarely seen before, no single lineage was responsible for symptomatic or asymptomatic infection – suggesting an important role for host factors. However, consistent with other countries, poultry-associated strains were a major source of infection. This genomic surveillance approach, that integrates microbial ecology with population based studies in humans and animals, has considerable potential for describing cryptic epidemiology in LMICs and will inform work to improve infant health worldwide.

## Introduction

The World Health Organization ranks diarrheal disease as the second most common cause of mortality among children under five years of age in low- and middle-income countries (LMICs), accounting for 10.6 million annual deaths in this age group [1,2]. *Campylobacter* is the most common cause of bacterial gastroenteritis in Europe and the USA, with even higher incidence in LMICs (up to 85% of children infected before 12 months [3]). However, *Campylobacter* infection is largely overlooked in LMICs for several reasons. Infection is thought to be sporadic so outbreaks are seldom recorded. *Campylobacter* are also more difficult to grow in the laboratory than many common enteric pathogens, so it is often not cultured even when present. These factors conspire such that the people at the greatest risk are the least studied.

In high-income countries, human campylobacteriosis is readily diagnosed as a disease associated with consumption of contaminated food, especially poultry [4,5], but the extremely high incidence in LMICs suggests different epidemiology. High exposure rates [6,7] and apparent endemism among young children [8–10] are a major concern, particularly as frequent or chronic (re)infection is linked to significant morbidity, growth faltering, cognitive impairment, and even death [11,12]. However, there is also evidence of common asymptomatic carriage among children in LMICs [7], a phenomenon that is not well understood. International studies have begun to quantify the causes of enteric infection in children [13–16] but campylobacteriosis surveillance programs remain uncommon and the strains responsible for disease are seldom characterized in LMICs [11,17–23]. Understanding the true disease burden requires not only incidence data, but also knowledge of variation in disease symptoms and the genotypes associated with asymptomatic and severe infection.

DNA-sequence-based strain characterization, typically of isolates from developed countries, has revealed considerable diversity within the major disease-causing *Campylobacter* species (*C. jejuni* and *C. coli*). This has allowed identification of the genotypes, and in some cases genes, linked with variation in disease symptoms and the source of infecting strains. For example, the identification of host-associated genetic variation [24] and the extent to which this segregates by host (host generalist and specialist genotypes) [25–27], means that human infection can be attributed to a specific reservoir source, when there is no human-to-human transmission [24,25,27–29]. Furthermore, in some cases it is possible to link particular genotypes to common disease sequelae [30–32] or severe infections [33–35], and identify locally [36–38] and globally distributed strains [39,40].

Among the most fundamental challenges in LMICs is to understand if disease severity and asymptomatic carriage are dictated by host factors, such as malnutrition [12], or the source and genotype of the infecting strain. In this study we address this as part of ongoing surveillance in Santa Clara, a semi-rural community near Iquitos in the Peruvian Amazon (**Figure 1A**). *C. jejuni* were isolated from individuals with varying disease severity, from no symptoms to severe infection, and the genomes were sequenced and contextualized within a global reference collection. Both, locally and globally disseminated genotypes were isolated from Peruvian children with a range of disease symptoms. Comparative genomics of isolates from symptomatic and asymptomatic individuals identified signatures of local diversification but little evidence of genetic elements specifically responsible for severe disease. Household crowding, poor sanitation, consumption of contaminated water and cohabitation with animals remain potential risks for local transmission, but poultry were revealed as an important infection reservoir based on source attribution analysis. This study provides a basis for considering complex transmission networks in LMICs and highlights the role of globally transmitted *Campylobacter* lineages.

**Figure 1.**
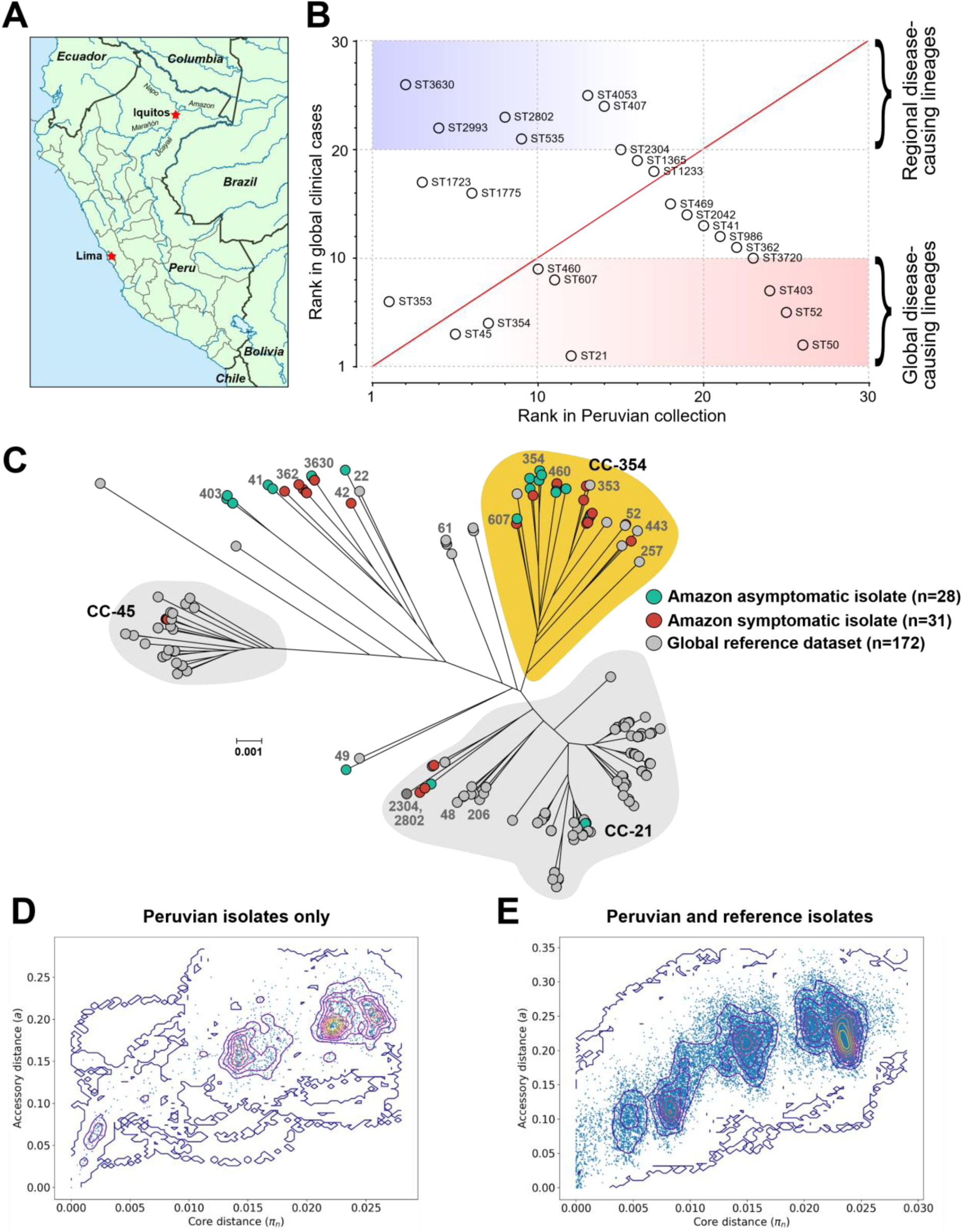
(**A**) Location of study site in Santa Clara, near Iquitos in Peru. (**B**) Sequence types (STs) of isolates collected from children in the Peruvian Amazon ranked according to the frequency in our local dataset and how often they have been sampled from human disease isolates (data from pubMLST; https://pubmlst.org/). (**C**) Population structure of *C. jejuni* isolates used in this study. All core (present in ≥95% of isolates) genes from the reference pan-genome list (2,348 genes) were used to build alignments of the Peruvian isolates (n=62) contextualized with 172 previously published genomes representing the known genetic diversity in *C. jejuni* (n=234, alignment: 720,853 bp. A maximum-likelihood phylogeny was constructed with IQ-TREE, using a GTR model and ultrafast bootstrapping (1,000 bootstraps; version 1.6.8) [55,56]. Scale bar represents genetic distance of 0.001. Leaves from asymptomatic Peruvian isolates are colored green; symptomatic Peruvian isolates are red; and isolates from the reference dataset are grey. Common STs and clonal complexes (CC), based on four or more shared alleles in seven MLST housekeeping genes, are annotated [60]. Interactive visualization is available on Microreact [57]: https://microreact.org/project/CampyPeruContext. (**D**) Pairwise core and accessory genome distances were compared using PopPunk for the Peruvian pediatric genomes only and (**E**) with the global reference dataset (version 1.1.4) [70].

## Methods

### Sampling and cohort information

Samples collected as part of a cohort study from Iquitos, in the Peruvian Amazon, between 2002 and 2006. In this age-stratified sample set of 442 children aged 0-5 years [7,13–15,41,42], children were visited 3 times weekly to form a continuous symptom history of childhood illnesses. Stool samples were collected quarterly from all children and in cases in which diarrhea was detected (92.3% of episodes detected by surveillance had a sample collected; **Table S1)**. Fecal samples were swabbed into Cary-Blair transport media, suspended in PBS, filtered through a 0.45 µm membrane and placed on a Columbia Blood Agar base (Oxoid) supplemented by 5% defibrinated sheep’s blood for 30 minutes prior to removal and streaking of filtrate. The Johns Hopkins Institutional Review Board provided ethical approval for the MAL-ED study in addition to respective partner institutions for each site, including Asociacion Benefica PRISMA, and the Regional Health Department of Loreto, Peru. Written consent was obtained from all participants.

### Bacterial isolate genome sequencing

Genomic DNA was extracted from 62 *C. jejuni* isolates and sequenced using an Illumina MiSeq benchtop sequencer (California, USA). Nextera XT libraries (Illumina, California, USA) were prepared and short paired-end reads (250 bp) were assembled *de novo* using Velvet (version 1.2.08) [43] with VelvetOptimiser (version 2.2.4). The average number of contiguous sequences (contigs) was 262 (range: 53–701) for an average total assembled sequence size of 1.55 Mbp (range: 1.37–1.70). The average N50 contig length (L50) was 14,577 (range: 3,794-55,912) and the average GC content was 30.8 % (range: 30.5-31.6). Short read data are available on the NCBI SRA, associated with BioProject PRJNA350267. Assembled genomes and supplementary material are available from FigShare (doi:10.6084/m9.figshare.10352375; individual accession numbers and assembled genome statistics in **Table S2**). Isolates were compared to a global reference dataset representing the genetic diversity of the species (n=164 isolates from eight countries and three continents) (**Table S3**)[26,36,44–47].

### Diarrheal disease severity

As part of the ongoing surveillance efforts, a questionnaire was completed three times per week to record diarrheal symptoms for all members of the cohort [7,13,14], generating a continual illness record for the surveillance period. *Campylobacter* isolated from patients that did not display any symptoms two days before or after collection of the stool sample were considered asymptomatic. Diarrhea was defined by three or more semi-liquid stools reported over a 24-hour period, with episodes separated by at least three symptom-free days. Diarrheal severity symptoms were catalogued and details recorded of any symptom, including the number of diarrheal episodes, hematochezia (blood in the stool), fever, incidence of vomiting and anorexia (**Table S1**)[48].

### Core genome genealogies

A reference pan-genome file was constructed by combining open reading frames identified by RAST [49,50] in all the Peruvian isolates and the *C. jejuni* NCTC 11168 reference strain to maintain locus nomenclature [51]. Gene orthologues (≥70% sequence similarity) were identified and duplicates removed (size: 2,045,739 bp; **Supplementary file S1**). Two alignment files were constructed from concatenated gene sequences of all core genes (found in ≥95 % isolates) from the reference pan-genome list using MAFFT [52] on a gene-by-gene basis [53,54]: one for the Peruvian isolates only (size: 772,794 bp; **Supplementary file S2**); and a second alignment containing the Peruvian isolates plus the global reference collection (size: 720,853 bp; **Supplementary file S3**). Maximum-likelihood phylogenies were constructed in IQ-TREE (version 1.6.8) using the GTR+F+I+G4 substitution model and ultra-fast bootstrapping (1,000 bootstraps)[55,56]; and visualized on Microreact [57]: Peru only (https://microreact.org/project/CampyPeruOnly); Peru and the global reference dataset (https://microreact.org/project/CampyPeruContext).

### Molecular typing and diversity estimates

Isolate genomes were archived in BIGSdb and MLST sequence types (STs) derived through BLAST comparison with the pubMLST database [58–60]. Capsule polysaccharide (CPS) and lipooligosaccharide (LOS) locus types of each *C. jejuni* isolate were characterized from their raw sequence data: short read sequences were mapped to known capsule and LOS locus types using BLAST as previously described [61,62]. Simpson’s index of diversity (with 95% confidence limits) was calculated for sequence types in the Peruvian and global reference datasets using the equation:

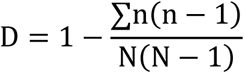

Where *n* is the number of isolates of each sequence type and *N* is the total number of isolates [55,63].

### Accessory genome characterization

The reference pan-genome list contained 2,348 genes, of which 1,321 genes were shared by all isolates (≥95 %) and defined as the core genome (**Table S4**). The accessory genomes of each isolate was characterized, including detection of antimicrobial resistance genes, putative virulence factors and known plasmid genes using ABRICATE (version 0.9.8) and the CARD, NCBI, ResFinder, VfDB and PlasmidFinder databases (10^th^ September, 2019 update; **Table S5** and summarized in **Table S6**) [64–69]. Pairwise core and accessory genome distances were compared using PopPunk (version 1.1.4). PopPUNK uses pairwise nucleotide k-mer comparisons to distinguish shared sequence and gene content to identify divergence of the accessory genome in relation to the core genome. A two-component Gaussian mixture model was used to construct a network to define clusters (Components: 43; Density: 0.1059; Transitivity: 0.8716; Score: 0.7793) [70].

### Source attribution

Sequence type (ST) and clonal complex (CC) ecological association were assigned based on previous publication and the relative abundance of STs among different host/sources within pubMLST (**Table S7**) [26,58]. Probabilistic assignment of the source host of infection was estimated using Structure v2.3.4, a Bayesian model-based clustering method designed to infer population structure and assign individuals to populations using multilocus genotype data [27,28,36,71,72]. In the absence of contemporaneous reservoir samples from Peru, we used a random selection of MLST profiles from pubMLST (n=1,229; ∼300 isolates per putative source reservoir; **Table S8**). A global genotype collection can be used for reservoir comparison as it is known that host-associated genetic variation transcends phylogeographic signatures [27]. MLST profiles of known providence were used to train the model (from 13 countries - 98% European; collected from 1996-2018). Isolates were grouped by source reservoir: chicken (denoting chicken carcass, meat or broiler environments), ruminant (cattle, sheep or goat feces, offal, or meat), wild birds (including starlings, ducks and geese) or other animal (as listed in pubMLST).

Self-assignment of a random subset of the comparison data set was conducted by removing a third of the isolates from each candidate population (n=388). Structure was run for 10,000 iterations following a burn-in period of 10,000 iterations using the no admixture model to assign individuals to putative populations. The assignment probability for each source was calculated for each isolate individually and isolates attributed to the putative origin population with the greatest attribution probability. We report an average self-assignment score of 61% (range 56.5-63.6%) following five independent estimations, consistent with other studies [27,28,73,74].

## Results

### Globally circulating disease genotypes are found in the Peruvian Amazon

We sequenced and characterized a collection of *C. jejuni* isolates (n=62) from a longitudinal cohort study of children under the age of 5 years sampled from diarrheal episodes and stools collected by protocol in the absence of diarrheal illness (**Figure 1A**). Isolate genotypes were compared with all genomes deposited in the pubMLST database (97,012 profiles, data accessed 17^th^ February, 2020) and ranked according to how frequently they were found associated with human disease (**Figure 1B**). Nearly half of the isolates (n=29, 47 %) were from common lineages, isolated many times before and recorded in pubMLST (>50 MLST profiles; **Figure 1B; Table S7**). Symptomatic (n=16; 52 % of disease isolates) and asymptomatic (n=12; 43 % of carriage isolates) isolates belonged to nine STs (eight CCs), including ST-353 (n=13), ST-45 (n=4), ST-354 (n=3), ST-607 (n=2), ST-460 (n=2), ST21 (CC21, n=1), ST50 (CC21, n=1), 52 (n=1) and ST-403 (n=1) (**Tables S6**). Of these common globally-distributed STs, represented by three or more isolates, only ST-45 was associated with disease - with 75 % of isolates (3 of 4) leading to symptomatic infection.

### Proliferation of globally rare genotypes in Peruvian Amazon children

The remaining 33 isolates (53 %) belonged to STs that are uncommon in the pubMLST database (<50 MLST profiles; **Figure 1B; Table S7**). This suggests that certain lineages that are rare in the UK and the USA may be more common among children in the Peruvian Amazon. Symptomatic (n=15; 48 % of disease isolates) and asymptomatic (n=16; 57 % of carriage isolates) isolates belonged to 17 STs (15 CCs), including ST-3630 (n=6), ST-1723 (n=5), ST-2993 (n=4), ST-1775 (n=3), ST-2802 (n=2), ST-535 (n=2), ST-362 (n=1), ST-3720 (n=1), ST-407 (n=1), ST-41 (n=1), ST-469 (n=1), ST-1233 (n=1), ST-1365 (n=1), ST-2042 (n=1), ST-2304 (n=1), ST-4053 (n=1) and ST-986 (n=1). Four of these rare STs were represented by three or more isolates: ST-3630 (4 of 6) and ST-2993 (CC362, 4 of 4) were predominantly symptomatic; while ST-1723 (CC354, 4 of 5) and ST-1775 (CC403, 3 of 3) were predominantly asymptomatic (**Table S6**).

All *C. jejuni* genomes (n=62) were compared to a global reference dataset representing known genetic diversity within *C. jejuni* (n=164 isolates from eight countries and three continents) using a maximum-likelihood phylogenetic tree (**Figure 1C**). Peruvian pediatric isolates did not cluster clearly by geography or disease severity. There was evidence that *C. jejuni* from children in the Peruvian Amazon represented a genetically diverse population. Specifically, there were 26 STs (19 CCs) among the Peruvian isolate collection, with a Simpson’s diversity index of 0.904 (95% CI: 0.863-0.946), compared to 50 STs (15 CCs) among the global collection of genomes (Simpson’s diversity index = 0.534, 95% CI: 0.453-0.615).

### Peruvian Amazon pediatric isolates have a local gene pool

While there were more STs in the Peruvian collection, there were fewer deep branching lineages compared to the global reference collection (**Figure 1DE**). This is not surprising as there were fewer samples in total and they came from a specific region and source (children). Discontinuous distribution of pairwise genomic distances in the Peruvian pediatric dataset is indicative of multiple genetically distinct clusters that are diverging in both core sequences and accessory gene content. Visualization of this clustering using the t-distributed stochastic neighbor embedding (t-SNE) projection of accessory distances tightly grouped the Peruvian isolates from the Amazon, while isolates from host generalist lineages in the global reference dataset (absent from the Peru dataset) were more loosely clustered (**Figure S1**). This provided evidence of increased horizontal gene transfer (HGT) among Peruvian isolates, compared to global isolate collection.

### Lineages associated with asymptomatic infection in Peruvian Amazon pediatric cases

Asymptomatic isolates and symptomatic isolates represented 17 STs (14 CCs) and 16 STs (14 CCs) respectively. Only 9 STs (8 CCs) contained a mixture of both disease etiologies. Of these common global STs represented by three or more isolates, only ST-45 was consistently associated with disease symptoms, with 75 % of isolates (3 of 4) leading to symptomatic infection (**Figure 2AB; Table S1**). Four rare STs: ST-3630 (4 of 6) and ST-2993 (CC362, 4 of 4) were predominantly symptomatic; while ST-1723 (CC354, 4 of 5) and ST-1775 (CC403, 3 of 3) were predominantly asymptomatic (**Figure 2AB; Table S1**).

**Figure 2.**
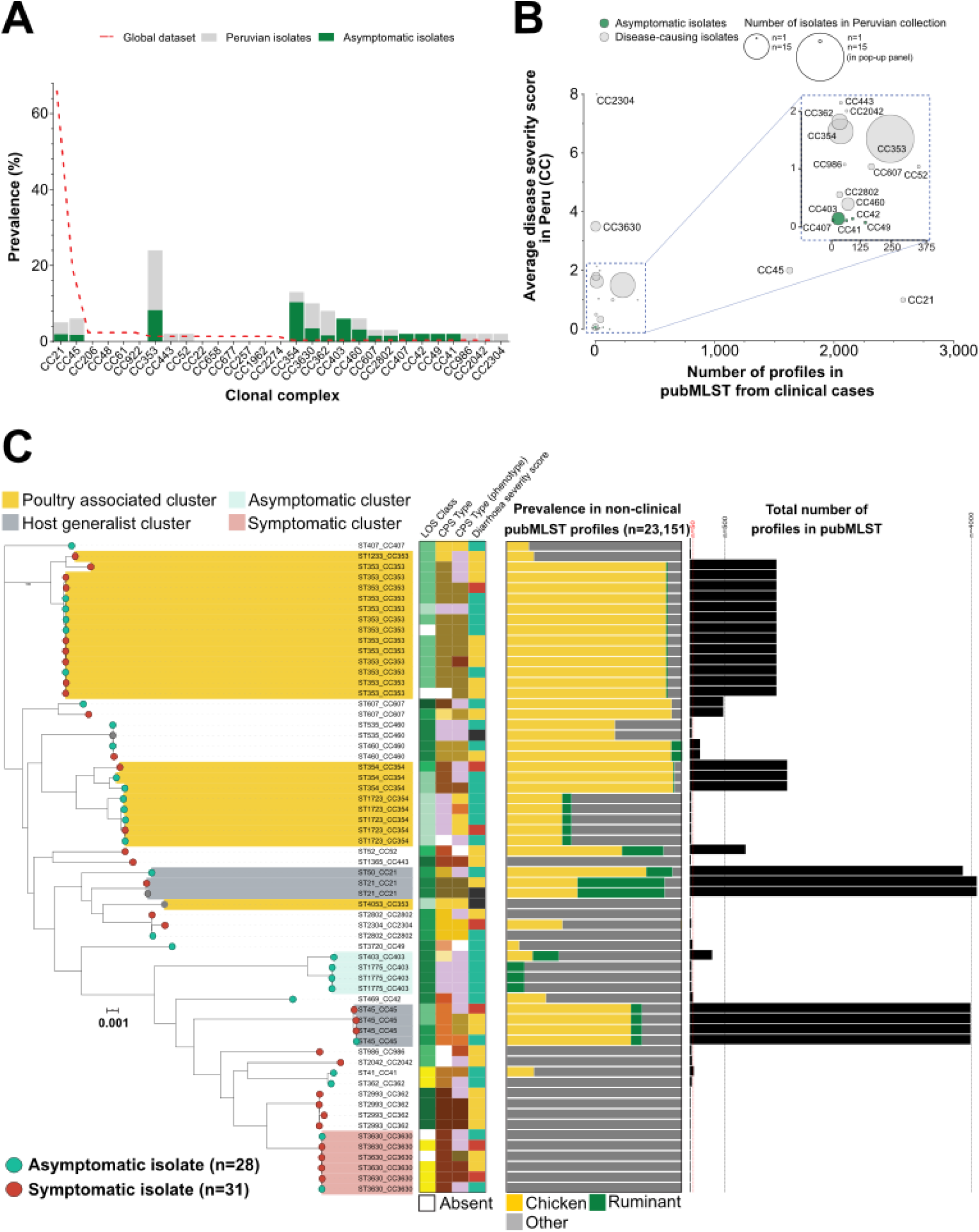
(**A**) Frequency of clonal complexes (CCs) identified among isolates collected from children in the Peruvian Amazon (grey bars) and the global reference dataset (red dotted line). Asymptomatic isolates are colored in green. (**B**) Average severity score of CCs represented by 3 or more genomes in our local dataset and how often they have previously been sampled from human disease (data from pubMLST; https://pubmlst.org/). Circle diameter represents how frequently they were sampled in our Peruvian Amazon pediatric collection. (**C**) A maximum-likelihood phylogeny was constructed with IQ-TREE, using a GTR model and ultrafast bootstrapping (1,000 bootstraps; version 1.6.8) [55,56] from an alignment of the Peruvian isolates only (n=62, alignment: 772,794 bp. Scale bar represents genetic distance of 0.001. Leaves from asymptomatic isolates are colored green and symptomatic isolates are red. The tree is annotated with lipooligosaccharide classes, capsular types and disease severity scores. Colored bar charts indicate the frequency with which the corresponding sequence type has been isolated from non-human hosts in pubMLST. Black bars indicate the overall frequency that the corresponding ST profile has been sampled before. Interactive visualization is available on Microreact [57]: https://microreact.org/project/CampyPeruOnly.

### Regional differences in accessory genome content

There was no difference in the mean genome size between symptomatic and asymptomatic isolates, but significant difference between the Peruvian Amazon pediatric population and the global reference dataset (ANOVA with Tukey’s multiple comparisons test, p-value <0.0001; **Figure S1AB**). This can partially be explained by a lack of isolates in the Peruvian pediatric collection from host generalist lineages, which tend to have larger genomes (ST-21 and ST-45 CCs; **Figure S1AB**), consistent with genome reduction being associated with increased host specialization [75,76]. As is typical of *Campylobacter* [35,44,54], the isolate collection included a large accessory genome (**Table S4**), with a little over half (56 %) the genes identified in the genomes of our 62 isolates from Peruvian children considered to be core (1,321 of 2,348 genes present in 95% of isolates). A large proportion of the accessory genome (446 genes, 43 % of the 1,027 accessory genes present in between 0 and 95 % of isolates) were present in less than 15 % of isolates.

Using the reference pan-genome list, genes that were core in the reference dataset were also present in the Peruvian pediatric dataset (average prevalence: 97.7 %) (**Figure S1C; Table S9**). All 29 of the NCTC11168 genes that were absent from Peruvian Amazon isolates (prevalence less than 5%) were found among genomes of isolates in the reference dataset (average prevalence: 43.0 %), with 21 specifically from the lipooligosaccharide (LOS) and capsular polysaccharide (CPS) loci. The LOS and CPS loci are highly variable in gene content [77–80] and this variability is reflected in the diversity of LOS and capsule types for the Peruvian isolates (n=14 LOS types; n=21 capsule types; **Figure S2**; **Table S6**). The most common LOS class locus was class H in 14 strains and 12/14 of these strains were poultry specialists and 10/14 strains were from symptomatic cases. LOS class B was present in 11 strains and only 2/11 were from symptomatic cases. There were four strains with LOS class A and all were from cases with symptomatic etiology and also possessed the HS:41 CPS locus. The most common CPS Penner type was HS:3 (n=10) and 70% of these strains were from symptomatic cases and all ten had LOS class H (**Table S6**).

### Poultry is the predominant source of infection in Peruvian Amazon children

STs were attributed to a putative host source based on their predominant sampling source in a global collection on pubMLST (**Table S7**). Isolates from poultry specialist lineages, including the globally disseminated ST-353, ST-354, ST-607 and ST-460, were the most common source of infection (n=32; **Figure 2C, Table S7**). Isolates from rare lineages, scarcely found outside human clinical cases (ST-3630, ST-2993, ST-2802, ST-986, ST41, ST362 and ST2402) were associated with the most severe symptoms. Poultry specialist and clinical specialist STs had average community diarrhea severity scores of 1.57 (n=30, max: 8) and 2.13 (n=16, max: 13), respectively. No isolates from ruminant-associated lineages caused any disease symptoms in this sample population, however the total number of isolates that putatively were from a ruminant background was small (n=5). Few isolates were isolated from the common generalist STs that dominate clinical collections in developed countries: ST-21 clonal complex (n=3) and ST-45 clonal complex (n=4). Quantitative source attribution estimated that 78.4 % (n=5, range 56.5 – 87.1 %) of the *C. jejuni* isolates emerged from chickens based on 5 different probability estimates (**Figure S3**).

## Discussion

Chronic diarrhea and malnutrition are major threats to children’s health worldwide. However, despite the high incidence of campylobacteriosis and reported differences in disease epidemiology, there is limited understanding *Campylobacter* in LMIC’s. By linking sequence data with detailed clinical records from the Peruvian Amazon pediatric cohort study we were able to show that variation in disease presentation was reflected in bacterial genomes, specifically the source and distribution (local and global) of infecting *C. jejuni* strains.

The Peruvian Amazon pediatric isolate collection comprised a diverse assemblage of STs, including common disease-causing lineages and regional STs, that have rarely been sampled in Europe and the USA [47,81]. Globalization of industrialized agriculture has dispersed livestock worldwide [82], broadening the geographical distribution of *C. jejuni*. We found evidence of this pervasive spread with two of the three most common strains isolated in the Peruvian Amazon belonging to the poultry-associated ST-353 and ST-354 complexes [47]. Quantitative source attribution also implicated chicken as the most likely source of infection, consistent with comparable studies in Europe (**Figure S3**) [27–29,73,83].

In contrast to the profusion of poultry-associated lineages, there was a striking paucity of host generalist ST-21 and ST-45 clonal complexes [40] that are among the most common disease-causing lineages in Europe and North America. This has previously been observed in another LMICs, with very few ST-21 complex isolates cultured in surveys from Africa, SE Asia and South America [84–88]. Ruminant specialist lineages were also rare among the Peruvian pediatric samples (6.1 %) and the most common cattle associated lineage (ST-61 complex [25]) was completely absent. This is clear evidence of different epidemiology in LMICs and potentially suggests different routes to human infection.

Asymptomatic *Campylobacter* carriage represents an alternative epidemiological context to that which has been the basis for most clinical studies [7,89,90]. *C. jejuni* is typically thought to cause transient infection with little opportunity for human-to-human transmission. This means that the human is an evolutionary dead end and the bacterium is unlikely to adapt to the human host. The high prevalence, regular reinfection and prolonged colonization periods in the Peruvian Amazon cohort study (and likely other LMICs) provide greater opportunity for human-to-human spread and adaptation to the host [91,92]. Some studies have attempted to identify signatures of human tropism, or even adaptation [93,94] and it remains possible that the some of the Peruvian STs that are rarely isolated from non-human infections (**Table S7**) could provide evidence of human adaptation.

One such candidate for human tropism in the Peruvian Amazon is the ST-403 complex (**Table S7**) [76]. None of the four ST-403 isolates we sampled were associated with diarrheal symptoms (**Table S1**), and according to many interpretations, attenuated virulence is often associated with long-term transmission [95]. This ST has also been sampled from human infections in the Dutch Antilles [96] and is a poor colonizer of avian hosts, typically lacking a gene cluster (*Cj1158-1159-1160*; **Figure S1C**; **Table S9**) [76] known to be important in chicken colonization [97]. However, not only was this gene cluster common in the Peruvian Amazon pediatric *C. jejuni* data but also there was no clear phylogenetic distinction between symptomatic and asymptomatic isolates, with multiple clonal complexes linked to asymptomatic carriage. While it remains possible that analysis of larger datasets will identify human adapted genomic signatures, our study suggests that host factors, such as cohabitation and poor sanitation, rather than the circulation of asymptomatic lineages, may be responsible for repeated or long-term infection.

While disease severity is not explained by specific lineage associations it remains possible that specific molecular variations mediate virulence in the Peruvian Amazon cohort. The intimate interaction of LOS and CPS with the host immune system means that the underling genes are a useful target for identifying genomic variation associated with asymptomatic carriage [61,98–100]. Hypervariable genes that are common in the reference dataset included several from the class C LOS and HS:2 CPS gene clusters (21 of 29 genes absent in ≥95 % Peruvian Amazon isolates), which are absent from the Peruvian Amazon pediatric isolates [62,101]. The LOS locus can be involved in the synthesis of LOS structures that mimic gangliosides, which play a role in the onset of several *Campylobacter* disease sequelae, including post-infectious neuropathies [76–80]. Although, there were no reports of these post-infectious neuropathies in any of these cases, there were 15 Peruvian isolates possessing LOS classes (A or B) that have been shown to be associated with Guillain-Barré and Miller syndromes [102–104]. Among these, all of the strains with LOS class A (n=4) were from symptomatic cases, while only 2 of 11 strains possessing LOS class B were from symptomatic cases. It should be noted that strains possessing LOS class B are not characterized by low virulence with strain 81-176 considered to be a highly virulent *C. jejuni* strain. Similarly, LOS classes that produce non-sialylated LOS also came from cases with differential etiology with 10 of 14 strains possessing class H from symptomatic cases and one of seven class K strains from symptomatic cases (**Table S6**).

Peruvian Amazon isolates were likely to have retained the ability to glycosylate flagella through genes contained in the O-linked glycosylation gene cluster (*Cj1293-1342c*), with each gene present in on average 73% (range 33.3 – 100%) of Peruvian Amazon isolates (**Table S9**). Large portions of the capsular polysaccharide (CPS) gene cluster appear absent from our local Peruvian Amazon isolates (*Cj1421c-Cj1441c*), however the flanking regions involved in capsule assembly and transport are highly conserved in our isolates (*kps* genes; **Table S9)**[77–80,105]. These differences are important to characterize and take into account during vaccine development for *Campylobacter*.

In conclusion, by contextualizing *C. jejuni* genomes from Peruvian Amazon children within a global reference collection and linking them to clinical data on varying disease symptoms and severity, we were able to identify local and globally distributed genotypes and determine the major source of infection (poultry). Furthermore, we show that common asymptomatic carriage is not the result of a single (or few) human adapted lineages suggesting an important role for host factor in long-term infections. Genomic surveillance integrating microbial ecology with population based studies in humans and animals, has considerable potential for describing cryptic epidemiology and untangling complex disease transmission networks in LMICs where interventions to reduce diarrheal disease are urgently needed.

## Data Availability

Short read data are available on the NCBI SRA, associated with BioProject PRJNA350267. Assembled genomes and supplementary material are available from FigShare (doi:10.6084/m9.figshare.10352375; individual accession numbers and assembled genome statistics in Table S2).

https://www.ncbi.nlm.nih.gov/bioproject/PRJNA350267

https://doi.org/10.6084/m9.figshare.10352375

## Abbreviations

LMIC: low- to middle-income country
GEMS: The Global Enteric Multicenter Study
MAL-ED: Malnutrition and Enteric Disease Study
MLST: multi-locus sequence typing
ST: sequence type
CC: clonal complex
LOS: lipooligosaccharide
AMR: antimicrobial resistance
CPS: capsular polysaccharide

## Supplementary materials

**(https://doi.org/10.6084/m9.figshare.10352375)**

**Supplementary Table S1:** Isolate list and disease severity scores

**Supplementary Table S2:** Assembly metrics and accession numbers

**Supplementary Table S3:** Global reference dataset details

**Supplementary Table S4:** Reference pan-genome gene presence

**Supplementary Table S5:** ABRICATE summary

**Supplementary Table S6:** Genome characterization

**Supplementary Table S7:** ST summary of pubMSLT

**Supplementary Table S8:** Source attribution dataset

**Supplementary Table S9:** Comparison of NCTC11168 gene presence

**Supplementary figure S1:**
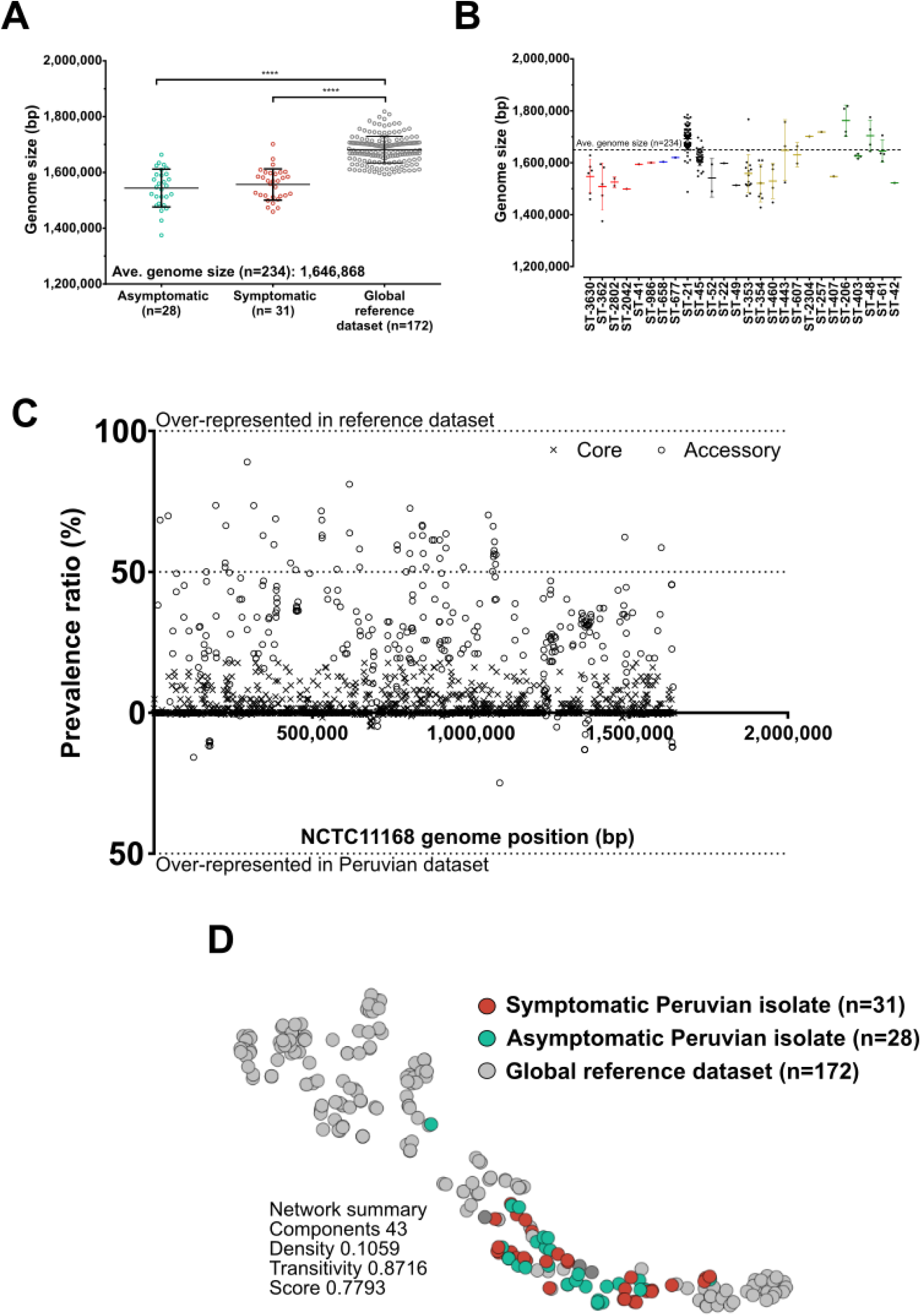
Genome size comparisons between (**A**) Asymptomatic (green) and symptomatic (red) Peruvian isolate genomes with the reference dataset (grey); and (**B**) all sequence types (ST) represented by 3 or more genomes in the dataset. Dotted line indicates the average genome size for all isolates in the dataset (1,646,868 bp). (**C**) Relative presence of all NCTC 11168 genes (n=1,623) in the Peruvian and reference datasets. Genes core and accessory in the reference dataset are indicated by (x) and (o), respectively. Genes present more often in one dataset compared to the other appear further from the mid-line. (**D**) Pairwise core and accessory genome distances were compared using PopPunk for the Peruvian genomes and full dataset (version 1.1.4) [70]. Clustering visualized using the t-distributed stochastic neighbor embedding (t-SNE) projection of accessory distances in microreact.

**Supplementary figure S2:**
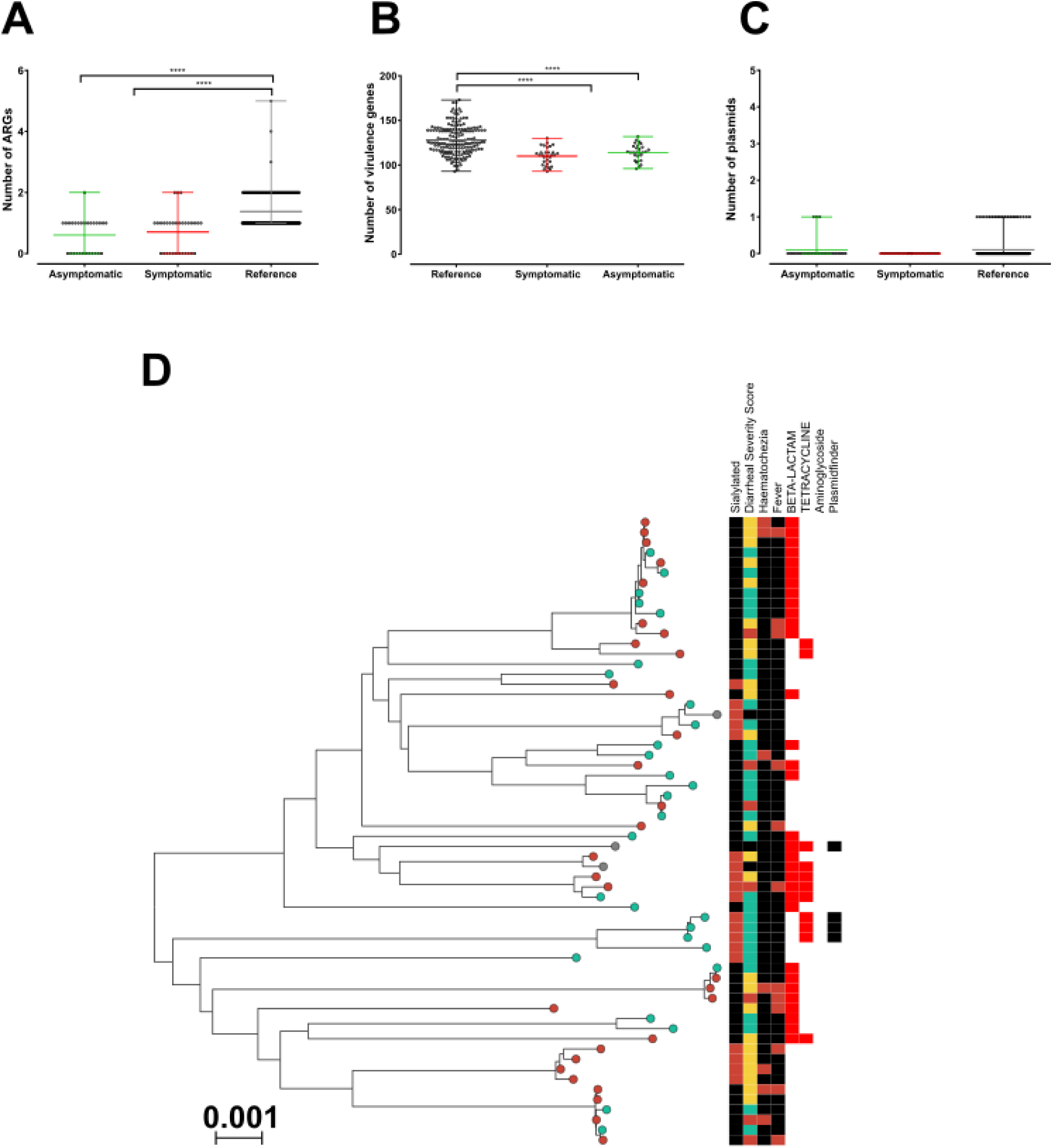
Number of (**A**) antimicrobial resistance genes (ARGs), (**B**) virulence genes and (**C**) predicted plasmids per isolate estimated using ABRICATE (version 0.9.8; [69]). (**D**) Maximum-likelihood phylogeny of the Peruvian isolates only. The tree is annotated with disease severity scores, the onset of specific symptoms (hematochezia and fever), presence of AMR genes (beta-lactams, tetracyclines or aminoglycosides), identified plasmids and sialylation prediction.

**Supplementary figure S3:**
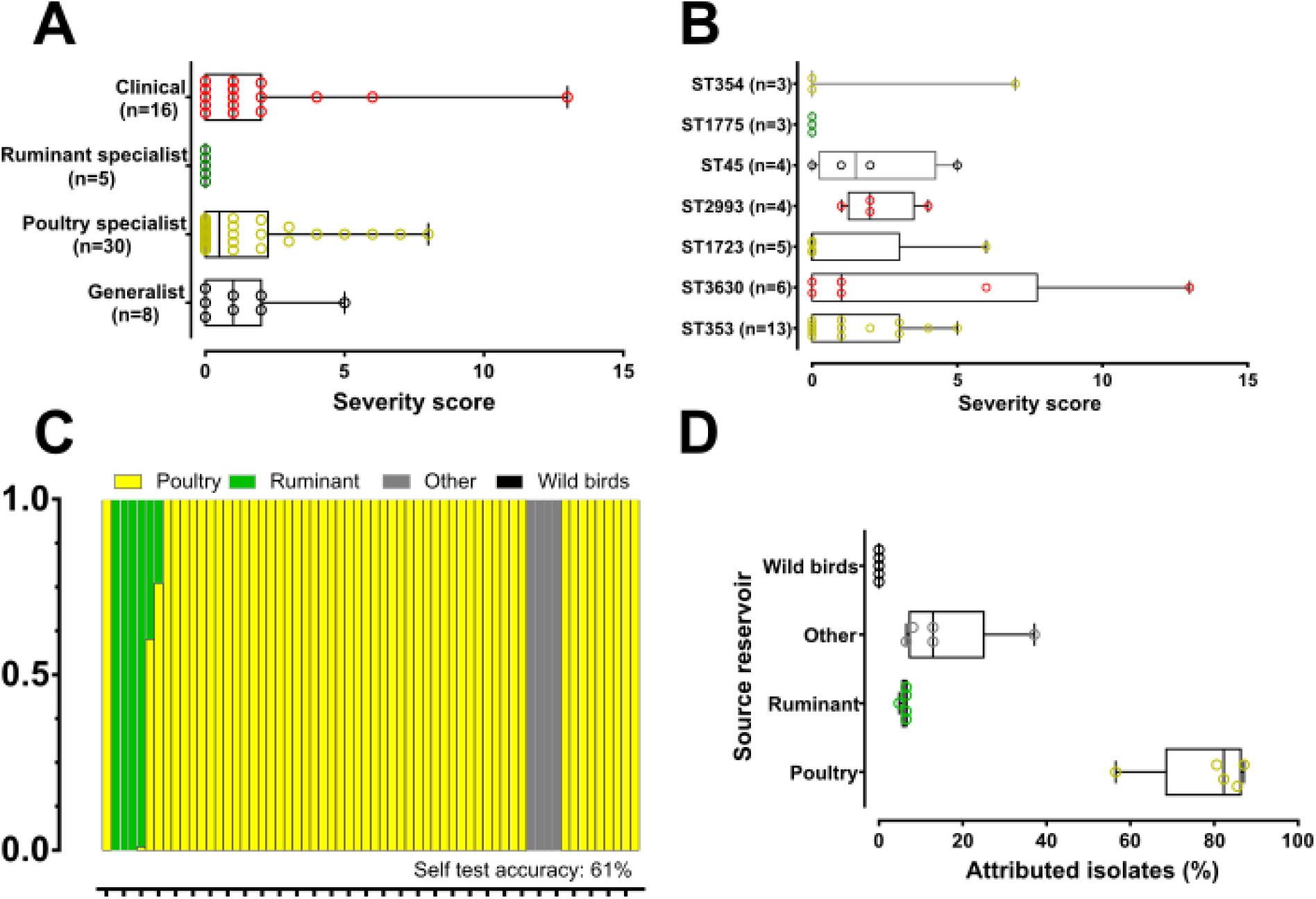
Average disease severity score by (**A**) isolate host ecology and (**B**) sequence type (represented by 3 or more isolates). (**C**) Representative source attribution of Peruvian pediatric isolates using the Bayesian clustering algorithm STRUCTURE (version v2.3.4, [71]). Each isolate is represented by a vertical bar colored by the estimated probability that it originated from putative source reservoirs (yellow: chicken; green: ruminant; black: wild bird and grey: other). (**D**) Summary box plots of predicted attribution of 62 Peruvian pediatric isolates following 5 independent estimations.

**Supplementary file 1:** Pan-genome

**Supplementary file 2:** Alignment – Peru isolates only

**Supplementary file 3:** Alignment – Peru plus context isolates.

## Contributors

Conceptualization: BP and SKS. Data Curation and Investigation: FS, RB, PY, MPO and MK led collection of the isolates. BP, SM and MDH sequenced the isolates. Formal Analysis: BP, FS, SM, SCB, GM, EM, JKC, KKC and CTP. Resources: BP, KAJ, MCJM and SKS. Original Draft Preparation: BP, CTP, MK, SKS. All authors read and approved the final manuscript.

## Acknowledgements

SKS is funded by the Medical Research Council (MR/L015080/1) and BBSRC (BB/P504750/1). Isolate sampling and collection were funded by the National Institute of Health (K01-TW05717) and MK received further support from the Bill & Melinda Gates Foundation (OPP1066146). Whole genome sequencing of the isolates was funded by an EPSRC Bridging the Gaps award (Swansea University). All high-performance computing was performed on MRC CLIMB, funded by the Medical Research Council (MR/L015080/1). Sequencing of global context isolates were previously funded by the Wellcome Trust (088786/C/09/Z). This publication made use of the PubMLST website (http://pubmlst.org/) developed by Keith Jolley and Martin Maiden at the University of Oxford and funded by the Wellcome Trust. No funding agency had any role in the study design, data collection and analysis, decision to publish, or preparation of the manuscript.

